# SARS-CoV-2 disinfection in aqueous solution by UV_222_ from a krypton chlorine excilamp

**DOI:** 10.1101/2021.02.19.21252101

**Authors:** Richard T. Robinson, Najmus Mahfooz, Oscar Rosas-Mejia, Yijing Liu, Natalie M. Hull

**Author notes:** Corresponding author: Natalie Hull, 2070 Neil Ave, Hitchcock 417C, Columbus, OH,43210.

## Abstract

There is an urgent need for evidence-based development and implementation of engineering controls to reduce transmission of SARS-CoV-2, the etiological agent of COVID-19. Ultraviolet (UV) light can inactivate coronaviruses, but the practicality of UV light as an engineering control in public spaces is limited by the hazardous nature of conventional UV lamps, which are Mercury (Hg)-based and emit a peak wavelength (254 nm) that penetrates human skin and is carcinogenic. Recent advances in the development and production of Krypton Chlorine (KrCl) excimer lamps hold promise in this regard, as these emit a shorter peak wavelength (222 nm) and are recently being produced to filter out emission above 240 nm. However, the disinfection kinetics of KrCl UV excimer lamps against SARS-CoV-2 are unknown. Here we provide the first dose response report for SARS-CoV-2 exposed to a commercial filtered KrCl excimer light source emitting primarily 222 nm UV light (UV_222_), using multiple assays of SARS-CoV-2 viability. Plaque infectivity assays demonstrate the pseudo-first order rate constant of SARS-CoV-2 reduction of infectivity to host cells to be 0.64 cm^2^/mJ (R^2^ = 0.95), which equates to a D_90_ (dose for 1 log_10_ or 90% inactivation) of 1.6 mJ/cm^2^. Through RT-qPCR assays targeting the nucleocapsid (N) gene with a short (<100 bp) and long (∼1000 bp) amplicon in samples immediately after UV_222_ exposure, the reduction of ability to amplify indicated an approximately 10% contribution of N gene damage to disinfection kinetics. Through ELISA assay targeting the N protein in samples immediately after UV_222_ exposure, we found no dose response of the ability to damage the N protein. In both qPCR assays and the ELISA assay of viral outgrowth supernatants collected 3 days after incubation of untreated and UV_222_ treated SARS-CoV-2, molecular damage rate constants were similar, but lower than disinfection rate constants. These data provide quantitative evidence for UV_222_ doses required to disinfect SARS-CoV-2 in aqueous solution that can be used to develop further understanding of disinfection in air, and to inform decisions about implementing UV_222_ for preventing transmission of COVID19.

**ABSTRACT ART / TOC GRAPHIC:** 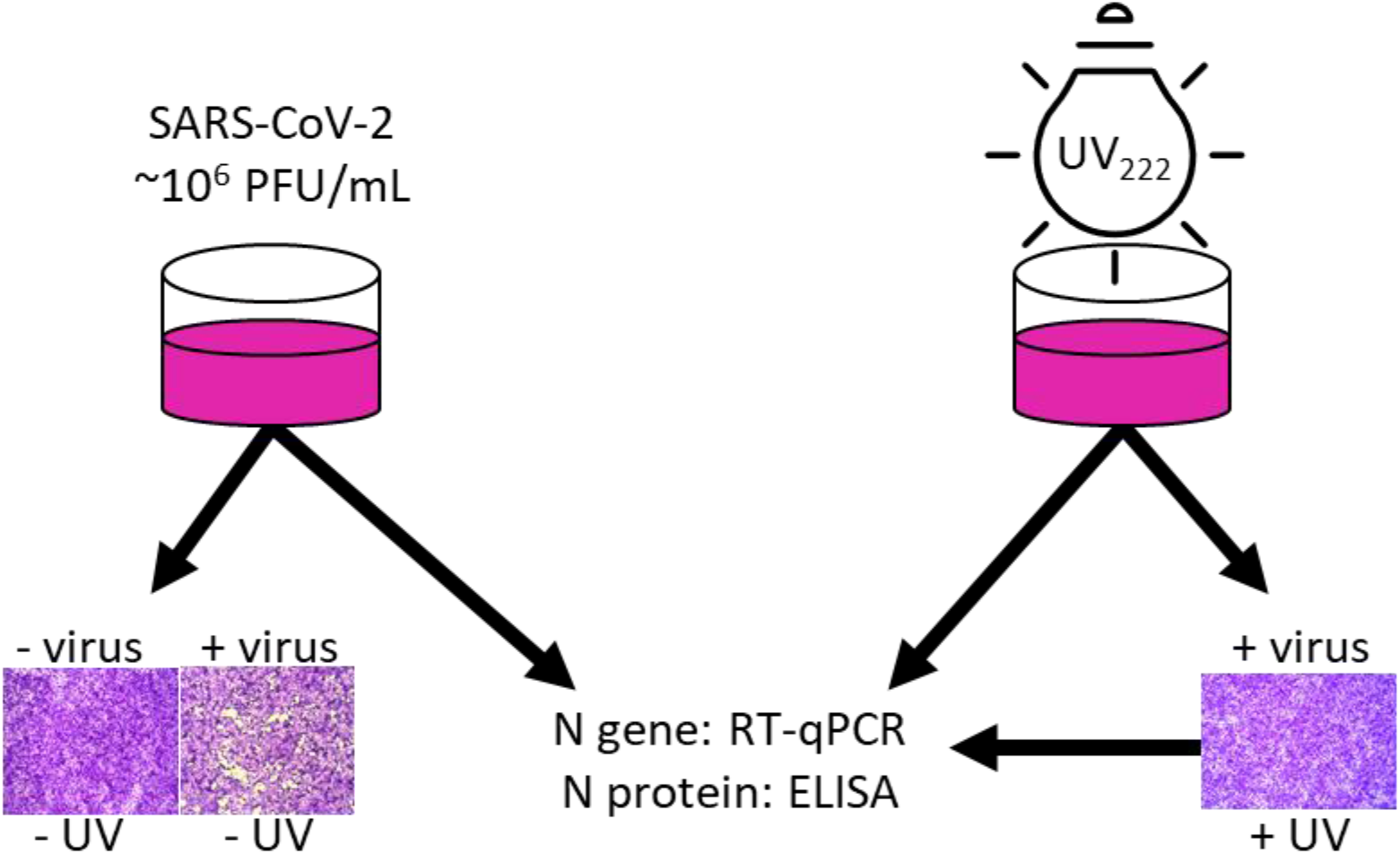

## INTRODUCTION

Severe acute respiratory syndrome coronavirus 2 (SARS-CoV-2) is the etiological agent of Coronavirus Disease 2019 (COVID-19), a recently emerged infectious disease with no cure. SARS-CoV-2 spreads primarily from person to person when mucous membranes (e.g., lungs, eyes) are exposed to airborne viruses that have been emitted by infected individuals in particles of various size^1,2^. Infection leads to a variable disease course affecting multiple organ systems (respiratory, cardiac, neurological and gastrointestinal); for this reason, the symptoms of COVID19 are variable and include asymptomatic infection, fever, cough, dyspnea, malaise, nausea, ageusia/anosmia, delirium and death. A number of antiviral and host-directed therapies have been or are being explored as COVID19 treatments, including low-dose radiation^3^, nucleoside analogs (e.g. remdesivir^4^, favipiravir^5^), hydroxychloroquine^6^, interferon beta^7^, convalescent plasma^8,9^, neutralizing monoclonal antibodies^10,11^, and anti-inflammatories such as dexamethasone^12^, IL6 inhibitors^13^, and JAK/STAT inhibitors^14^. Prophylactic vaccines against the SARS-CoV-2 Spike protein have also recently become available^15^. These treatments and vaccines are causes for optimism during the current COVID19 pandemic, which to date has killed nearly 2 million individuals; however, even after vaccines become widely available, social distancing, face masks and other engineering solutions that limit transmission will continue to be needed in the foreseeable future for this and other emerging infectious diseases^16^.

Ultraviolet (UV) irradiation is an effective means of inactivating a number of respiratory viruses, including human coronavirus OC43 (HCoV-OC43, a cause of the common cold^17^) and SARS-CoV (etiological agent of the 2002 SARS epidemic^18–20^). UV is commonly applied for upper room air disinfection, in HVAC systems, and in free-standing air and surface purifiers. The feasibility of using UV on a widespread and evidence-based level to minimize transmission of SARS-CoV-2, however, is currently limited by two reasons: (1) conventional mercury-based low pressure UV lamps are impractical in many settings as they are hazardous to human health (the 254 nm wavelength emission causes skin cancer^21^ and cataracts^22^) and the environment (mercury from breaking fragile quartz lamp bulbs is toxic^23^), (2) the UV dose response kinetics needed to inactivate SARS-CoV-2 are unknown. Should these two challenges be overcome, the use of UV to inactivate SARS-CoV-2 in environments with high potential for transmission (e.g. congregate care facilities, convalescent patient homes, hospital waiting rooms, airplane cabins) would be a practical and readily deployed engineering solution to augment current prophylactic measures (social distancing, face masks, vaccines). Due to a surge in interest and application of UV in various public settings, there is an urgent need to understand the dose response kinetics of SARS-CoV-2 to UV radiation to inform decisions which balance the risk to eyes and skin from UV exposure with the risk of infection from virus transmission.

Here we demonstrate the dose response kinetics of SARS-CoV-2 in liquid after exposure to primarily 222 nm UV light emitted by a krypton-chlorine (KrCl) excimer lamp (excilamp) filtered to reduce transmission of more harmful wavelengths > 240 nm. The lower wavelength emission (222 nm) is neither carcinogenic in human skin models or rodents^24^, nor causes acute corneal damage in rodents^25^. Additionally, the 222 nm wavelength emitted by KrCl excilamps is inherently more effective at disinfection^26^, nucleic acid damage^27^, and protein damage^28,29^ than 254 nm emitted by low pressure mercury lamps due to greater absorbance of target biomolecules at lower wavelengths. Krypton and chlorine in KrCl excilamps are much less toxic than mercury, and KrCl excilamps have already been shown to be competitive in terms of electrical efficiency with mercury lamps that have many more years of product development and optimization^30^. Our results demonstrate that when an aqueous solution of pathogenic SARS-CoV-2 is exposed to UV_222_ light emitted by a Kr-Cl excilamp, its infectivity and integrity is attenuated in a UV dose-dependent manner, as measured by culture and molecular assays. These first UV_222_ disinfection dose responses demonstrate the feasibility of UV as an approach to inactivate SARS-CoV-2.

## METHODS

### SARS-CoV-2 culture

SARS-CoV-2, Isolate USA-WA1/2020, was obtained from Biodefense and Emerging Infections Research Resources Repository (BEI Resources, Batch # 70034262) and stored and cultured in the Ohio State University Biosafety Level 3 laboratory (IBC Protocol # 2020R00000046). The viral stock used in this study was established by thawing the Batch, diluting it 1:10,000 into incomplete DMEM (Gibco Cat# 11995-065, supplemented with 4.5 g/L D-glucose, 110 mg/L sodium pyruvate), and adding it to T175 flasks of confluent Vero cells (ATCC clone E6) for a one hour incubation period (37°C, 5% CO_2_), after which the supernatant was removed and replaced with complete DMEM (cDMEM; DMEM as above plus 4% heat-inactivated fetal bovine serum). These T175 flasks were incubated for 3 days (37°C, 5% CO_2_) to propagate infectious virus. At the end of this period, visual inspection of the flasks under a light microscope demonstrated that the nearly all Vero cells were dead. The supernatants in each of the T175 flasks were presumed to contain infectious virus at this point, were carefully transferred and combined into a 50mL conical, centrifuged at low speed to remove cell debris, aliquoted into microcentrifuge tubes, frozen and stored at −80°C. The live virus titer in frozen aliquots was determined to be ∼10^7^ plaque forming unit (PFU) per mL using a modified version of plaque assay developed by the Diamond laboratory^31^ and described below.

### UV Dose Calculations

The UV_222_ light source (USHIO Care222^®^) is a KrCl excilamp that is optically filtered to reduce emission > 240 nm. The UV source was turned on to warm up for 15 minutes before any irradiance or spectral measurements or irradiations. Standardized procedures were followed for carrying out quasi-collimated beam disinfection studies^32^ and calculating polychromatic UV doses^33^. The emission spectrum of the UV_222_ source was measured using a NIST-traceable calibrated Ocean Optics HDX UV-Vis spectroradiometer with an extreme solarization resistant 455 µ fiber and Spectralon diffusing cosine corrector detector. Raw spectral data from the OceanView software was interpolated to integer wavelengths using the FORECAST function in Microsoft Excel and relativized to peak emission at 222 nm for use in dose calculations (Figures 1 and S1). Total incident UV-C irradiance was measured using an International Light Technologies (ILT) 2400 radiometer with a SED 220/U solar blind detector, W Quartz wide eye diffuser for cosine correction, and peak irradiance response NIST-traceable calibration. For irradiance measurement, the peak wavelength calibration value was input manually as the radiometer factor. The incident irradiance was measured with the detection plane of the radiometer centered at the height and location of the sample surface during UV exposures, and corrected for several factors to determine the average irradiance through the sample depth. Spatial nonuniformity of emission was accounted for each test by measuring irradiance at 0.5 cm increments from the center to the edge of the petri dish and relativized to determine a petri factor, which was always > 0.9. The typical detector spectral response was obtained from ILT and used to calculate the radiometer factor integrated over the lamp emission, which was 0.9971. As previously^34^, the reflection factor for water at the 222 nm peak wavelength was assumed to be 0.9726. The divergence factor was determined each experiment day by accounting for the distance between the lamp and the sample surface, and the sample depth and was always > 0.9. The water factor was determined each sample day by the ratio between the incident irradiance and the average irradiance integrated through the sample depth after wavelength-specific absorption. The UV-vis absorbance of virus working stocks (prepared fresh for each test) was measured in the biosafety cabinet using a Nanodrop™ One^C^ spectrophotometer via the microvolume pedestal for wavelengths 200 −295 nm and the 1 cm quartz cuvette for wavelengths above 195 nm. Working stock absorbance spectra for each test are shown in Figures 1 and S1. After these adjustments to incident irradiance in the center of the sample, the average irradiance was used to calculate exposure times (max: 15 minutes; min: 15 seconds) for pre-determined UV doses (0-40 mJ/cm^2^) (summarized in Supplementary Table S1).

**Figure 1:**
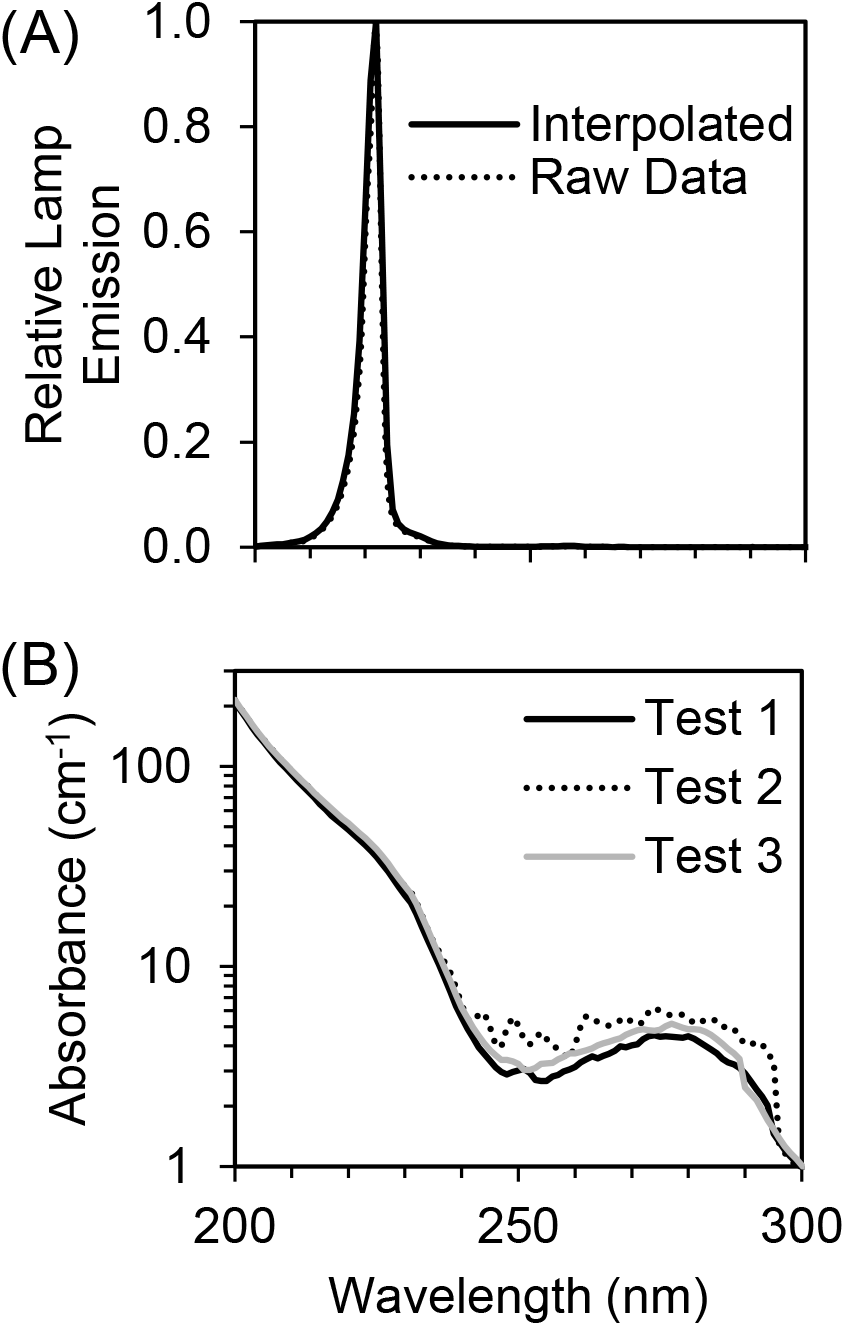
(A) The raw spectral emission from 200 - 300 nm of the filtered KrCl excilamp (USHIO Care222^®^) was interpolated and relativized to the peak emission at 222 nm for use in UV dose calculations. (B) The absorbance spectrum from 200 - 300 nm of SARS-CoV-2 at ∼10^5^ PFU/mL in cDMEM was measured for each of three biologically independent Tests for use in UV dose calculations. Expanded emission and absorbance spectra from 200 - 800 nm are shown in Supplementary Figure S1.

### UV Treatment

All UV measurements, sample preparation, UV treatments, and subsequent handling of treated samples were performed in a biosafety cabinet. On the day of each three biologically independent tests while the UV source warmed up and measurements were taken for dose calculations, aliquots of SARS-CoV-2 (previously tittered at 10^7^ PFU/mL) were diluted in cDMEM to make a “working stock solution” with a target titer of 10^5^ PFU/mL. For each UV dose tested, 3 mL of the working stock solution was pipetted into a 3.7 cm^2^ area and 3.5 cm diameter polystyrene tissue culture dish (VWR Catalog # 82050-538) with a sterile Teflon-coated micro stir bar (VWR Catalog # 58948-353) and positioned under the UV light on a small stir plate to achieve quiescent mixing while blocking the UV light with a shutter. After removing the tissue culture dish lid, the shutter was removed to expose the sample to UV light for the calculated exposure time corresponding to the pre-determined UV dose before replacing the aperture to end the UV exposure. Immediately afterwards, the treated media was transferred to a sterile 15 mL polypropylene centrifuge tube (VWR) and used for the assays described below. Working stocks for untreated samples were placed on the stir plate for a representative amount of time with the lamp off before transfer to centrifuge tube (0 mJ/cm^2^).

### SARS-CoV-2 plaque assay

Plaque assays were used to determine PFU/mL of samples before UV treatment (0 mJ/cm^2^) and after UV treatment (all other UV doses). The plaque assay used for this study is a modification of that which was originally developed and reported by Case et al,^31^ and is listed here as STEPS 1-5. (**STEP 1**) At least 18 hours prior to the assay, 12-well plates were seeded with a sufficient number of Vero cells so that each well was confluent by the assay start; plates were incubated overnight at 37°C. (**STEP 2**) On the day of the assay, serial dilutions of virus-containing media (e.g UV treated virus samples) were prepared in cDMEM (1:10^1^, 1:10^2^, 1:10^3^, 1:10^4^) and warmed to 37°C. (**STEP 3**) Media from each well of the 12-well plate was gently removed via pipette and replaced with 500uL of each virus serial dilution, the volume pipetted down the side of the well so as not to disturb the Vero cell monolayer. (**STEP 4**) The plate was incubated for one hour at 37°C, 5% CO_2_. (**STEP 5**) During that infection incubation period, a solution comprising a 1:0.7 mixture of cDMEM and 2% methylcellulose (viscosity: 4000 cP) was freshly made and warmed to 37°C in a water bath. After the one hour infection incubation period, the supernatant was removed from each well and replaced with 1 mL of the warmed cDMEM/methylcellulose mixture. (**STEP 6**) The culture plate was then returned to the incubator and left undisturbed for 3 days. On the final day, cDMEM/methylcellulose mixture was removed from each well, cells were fixed with 4% para-formaldehyde in PBS (20 minutes, room temperature), washed with PBS and stained with 0.05% crystal violet (in 20% methanol). After rinsing plates with distilled water, plates were dried and plaques were counted under a light microscope at 20X magnification.

### SARS-CoV-2 outgrowth assay

The virus outgrowth assay used for this study is identical to the plaque assay described above, with the exception that after **STEP 4** the virus laden media was replaced with 1 mL of warm cDMEM (instead of a cDMEM/methylcellulose mixture). Afterwards, the culture plate was returned to the incubator and left undisturbed for 3 days. On the final day, the cell supernatants of each well were collected, transferred into a microcentrifuge tube, centrifuged at low speed to remove cell debris (1,000 × *g*, 10 min), aliquoted into microcentrifuge tubes, frozen and stored at −80°C. Aliquots were subsequently used for quantitative real time PCR (qRT-PCR) measurement of SARS-CoV-2 nucleocapsid (N) gene copies, as well as ELISA determination of SARS-CoV-2 N protein concentrations.

### SARS-CoV-2 N gene quantitation -N1 primer set

Quantitative PCR (qPCR) was used to quantify the SARS-CoV-2 N gene directly in RNA extracts of samples before UV treatment (0 mJ/cm^2^) and after UV treatment (all other UV doses), and in RNA extracts of cell supernatant aliquots from outgrowth assays. RNA was extracted from samples using the QIAamp Viral RNA method (Qiagen), and converted to cDNA using the SuperScript IV first strand synthesis method with random hexamer primers (Invitrogen). cDNA was subsequently amplified with the “N1 primer set” and associated PCR conditions that were originally developed by the Centers for Disease Control^35^. These primers are specific to nucleotides 13-85 of the N gene (NCBI Ref Seq NC_045512.2) and generate a short (72 nt) amplicon: 2019-nCoV_N1-F (forward) primer, 5’-GACCCCAAAATCAGCGAAAT-3’; 2019-nCoV_N1-R (reverse) primer, 5’-TCTGGTTACTGCCAGTTGAATCTG-3’. cDNA was PCR-amplified in a quantitative PCR (q-PCR) assay comprising 1X TaqMan Universal PCR Master Mix (Applied Biosystems), the N1 forward/reverse primers described above (final concentration: 500 nM) and a fluorophore-conjugated N1 TaqMan probe (5’-FAM-ACCCCGCATTACGTTTGGTGGACC-BHQ1-3’; final concentration 125 nM). q-PCR assays were run on a BioRad CFX Connect Real Time PCR system to determine C_T_ values from samples and standards. A standard curve was generated for the N1 primer set by running serial dilutions on each plate of *in vitro* transcribed RNA converted to cDNA relating N gene copy numbers to C_T_ values. To generate this standard, RNA was extracted from an aliquot of our SARS-CoV-2 stock and converted to cDNA before amplification of the N gene using the N1 primer set as described above. The amplicon was visualized by agarose gel electrophoresis, gel extracted and cloned/ligated into the plasmid vector pCR II-TOPO (Invitrogen), downstream of the T7 promoter. Ligation products were transformed into *E. coli*, and mini-preps of randomly selected colonies were screened via PCR for the presence of insert. A single clone was then used to produce *in vitro* transcribed (IVT) N gene RNA—a reagent necessary for accurate gene copy number measurement—using the HiScribe T7 Quick High Yield RNA Synthesis method (New England Biolabs). After treating the IVT RNA with DNase and performing a cleanup reaction, the RNA concentration was determined via Nanodrop. The copies of single stranded N gene RNA transcripts per µL was determined by the following equation: [RNA concentration (Nanodrop measurement, ng/µL) × the Avogadro number (6.02 × 10^23^)] / [Predicted molecular weight of transcript (23 kDa) × 10^9^]. Serial dilutions of IVT RNA were made (range: 10^13^→ 10^−1^ copies/μL), converted to cDNA as above and used as standards in the N gene copy number assay described above.

### SARS-CoV-2 N gene quantitation -N1-2 primer set

qPCR was used to quantify the SARS-CoV-2 N gene in RNA extracts of samples of working stocks before UV treatment (0 mJ/cm^2^) and immediately after UV treatment (all other UV doses). RNA was extracted from samples and converted to cDNA as described above. cDNA was subsequently quantified using a combination of the CDC 2019 N1 and N2 primer sets to generate a long (944 nt) amplicon: 2019-nCoV_N1-F (forward) primer, 5’-GACCCCAAAATCAGCGAAAT-3’; 2019-nCoV_N2-R (reverse) primer, 5’-GCGCGACATTCCGAAGAA-3’. Primers were obtained from IDT and final concentrations were 500 nM, in 10 μL SsoFast EvaGreen Supermix (BIO-RAD) and 7.75 μL nuclease free water (Fisher Scientific) and 2 μL cDNA template. Reactions with total volume of 20 μL were run in at least technical duplicate on an Applied Biosystems QuantStudio 7 Real-Time PCR system to determine C_T_ values from samples and standards. For the N1-2 primer set, the standard consisted of serial dilutions of the double stranded DNA control plasmid of the complete N gene (2019-nCoV_N_Positive Control, IDT).

### SARS-CoV-2 N protein ELISA

The concentration of N protein in outgrowth assay supernatants was determined using the SARS-CoV-2 Antigen Quantitative Assay Kit (ELISA) method (ADS Biotec). Manufacturer-provided calibration controls were used to establish a standard curve related N protein concentration to sample absorbance (wavelength: 450 nm). Values outside the standard curve were diluted further and rerun as appropriate. The positive signal for SARS-CoV-2 was 2.7 × 10^5^ ±9.8 × 10^4^ pg/mL in untreated virus samples at Day 0 and 1.4×10^8^ ±3.0 × 10^8^ pg/mL in cell culture supernatants incubated with untreated virus samples at Day 3. No N protein was detected in negative control cell culture supernatants that were incubated without virus samples.

### Graphing and Statistics

Graphs were prepared using either GraphPad Prism or Microsoft Excel programs; statistical analyses (including regression using the data analysis add-in to determine standard error of regression coefficients) were performed using these programs’ bundled software. Log_10_ Reduction (LR) was calculated as log_10_(N_o_/N), where N was viral PFU/mL in the plaque assay, N gene copies/µL in qPCR assays for either the short N1 amplicon or the long N1-2 amplicon, or N protein concentration in pg//mL in the ELISA assay after exposure to a given UV_222_ dose, and N_o_ was the initial concentration. The level of replication in this study was three biologically independent tests, with at least technical duplicates for each assay.

## RESULTS

### SARS-CoV-2 Infectivity Response to UV_222_

Viral infectivity UV_222_ dose response was characterized by exponential decay kinetics (Figure 2). At a mean initial viral titer of 6.51×10^4^ PFU/mL, the pseudo first order rate constant for viral disinfection was −1.48 cm^2^/mJ (R^2^ = 0.89). When expressed as LR of viral infectivity after exposure to a given UV dose, the linear rate constant was 0.64 cm^2^/mJ (R^2^ = 0.95), which equates to a D_90_ (dose for 1 log_10_ or 90% inactivation) = 1.6 mJ/cm^2^. Doses ranges and initial Vero cell confluence were only sufficient in the Test 3 experimental replicate to quantify a dose response. However, in Test 2, the mean initial viral titer of 3.54×10^4^ PFU/mL in untreated samples was reduced to below detection by the first dose tested of 10 mJ/cm^2^, equivalent to a LR of at least 4.25 logs. These results were also consistent with qualitative results from Test 1, where Vero cells appeared mostly dead in the untreated samples, appeared increasingly healthy through doses 0.7 and 1.4 mJ/cm^2^, and appeared healthy at doses above 2 mJ/cm^2^.

**Figure 2:**
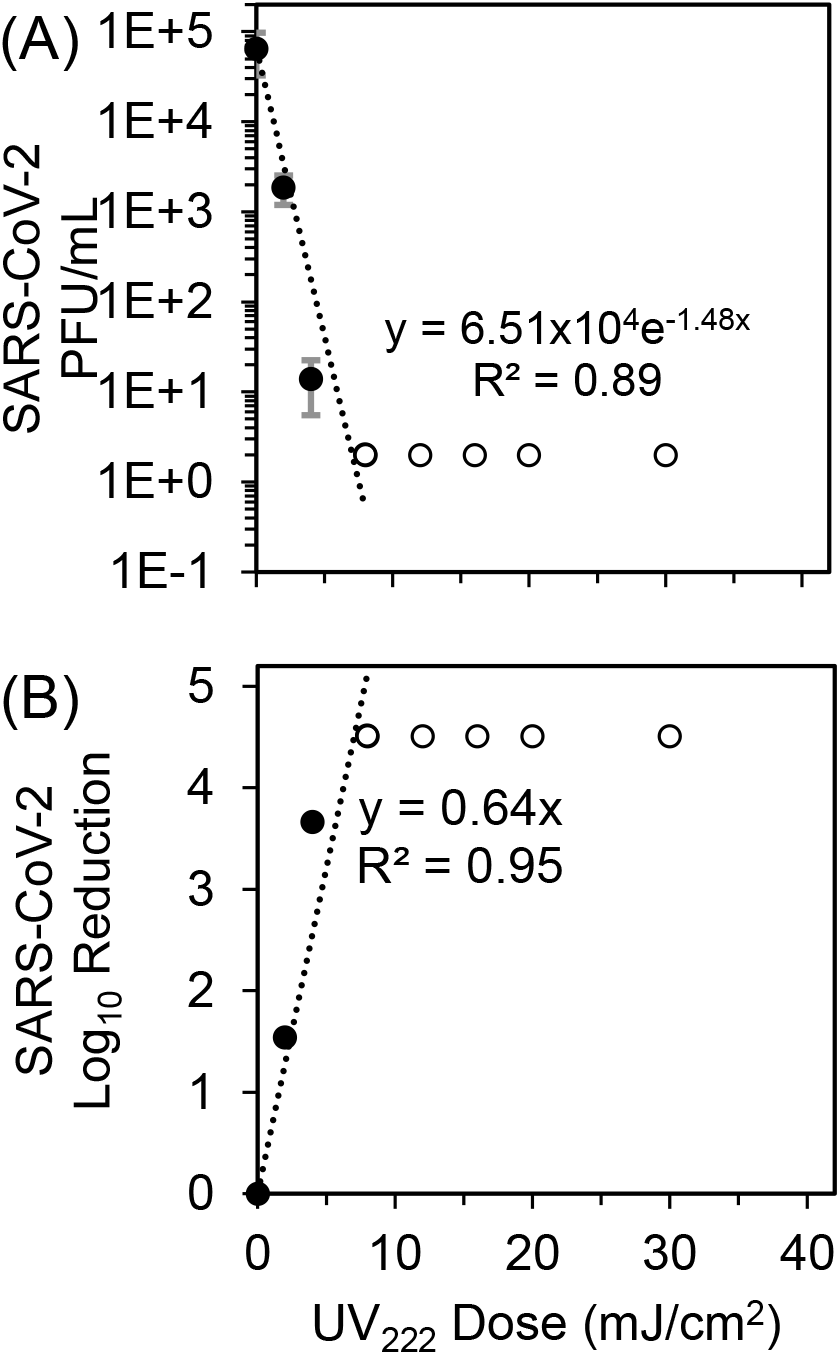
(A) SARS-CoV-2 titers measured by plaque assay 3 days after sample exposure to each UV_222_ dose (dark circles) were fit with an exponential model starting at the mean initial (0 mJ/cm^2^) viral titer of 6.51×10^4^ PFU/mL through responses up to and including 8 mJ/cm^2^ where PFU/mL first dropped below the assay detection limit (DL) of 2 PFU/mL (hollow circles). Error bars represent standard deviation of at least two technical replicates. (B) SARS-CoV-2 log_10_ reductions (LR) of viral titers after exposure to each UV_222_ dose (dark circles) were fit with a linear model forced through the origin at 0 mJ/cm^2^ through responses up to and including 8 mJ/cm^2^ where LR first exceeded the DL of 4.51 logs (hollow circles).

### SARS-CoV-2 N Gene and Protein Response to UV_222_

For the short amplicon spanning the N1 region of the N gene (CDC 2019), viral RNA damage in response to UV_222_ immediately after treatment was also characterized by exponential decay kinetics (Figure 3A). When expressed as LR of N1 copies/µL in qPCR reactions after exposure to a given UV dose, the linear rate constant was 0.069 ±0.005 cm^2^/mJ (slope ±standard error, R^2^ = 0.92). The N1 dose response was modeled using the linear region between 0 - 20 mJ/cm^2^ to avoid tailing in the dose response. When including only doses up to 10mJ/cm^2^ as for the plaque assay, the slope and R^2^ of the N1 gene damage dose response was the same as for doses up to 20 mJ/cm^2^. Compared with the LR rate constant for of SARS-CoV-2 infectivity measured by plaque assay, the LR rate constant of N gene damage measured by N1 qPCR was approximately 10-fold lower. Across all tests, the positive signal for SARS-CoV-2 in the N1 assay was 10.75 ± 0.25 log_10_ copies/μL in cell cultures infected with untreated virus (0 mJ/cm^2^), 4.89 ± 0.86 log_10_ copies/μL in uninfected cell culture supernatants, 5.49 log_10_ copies/μL in RNA extraction negative control, 3.36 ± 0.24 log_10_ copies/μL in no template RT-qPCR reaction controls (concentration data and standard curves shown in Supplementary Figures S2 and S3). Despite this background signal, dose responses were still discernable. For the N1 dose response after 3 days in the outgrowth assay for doses up to 0 - 20 mJ/cm^2^, the linear rate constant was 0.260 ± 0.036 cm^2^/mJ (slope ± standard error, R^2^ = 0.76). Although a positive dose response was apparent and the slope was closer to the plaque assay (indicating better ability to predict plaque assay dose response with combined cell culture with qPCR), the increased variability introduced by cell culture decreased the strength of the regression.

**Figure 3:**
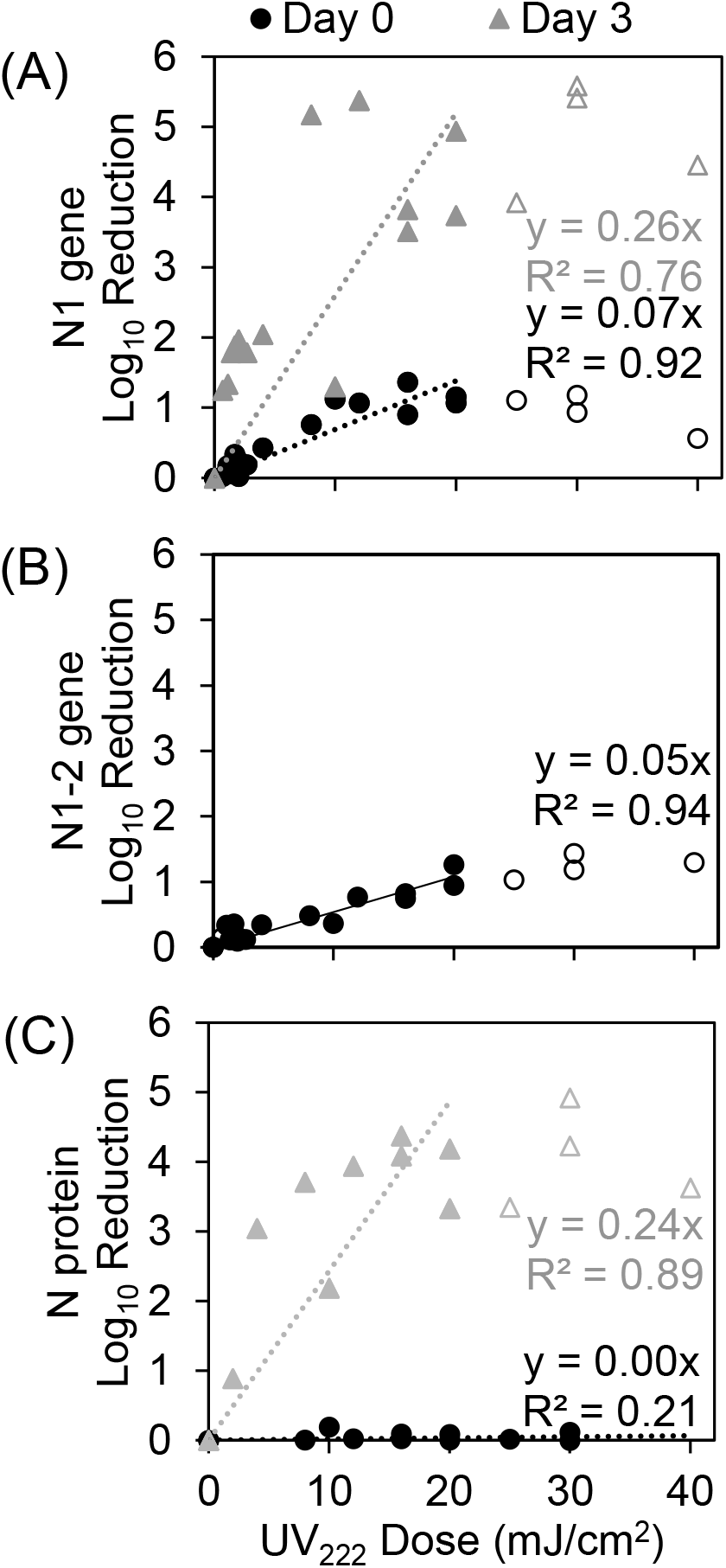
(A) SARS-CoV-2 N gene damage immediately after UV treatment (Day 0) and after incubation of samples with host cells (Day 3) expressed as log_10_ reduction of N1 (short amplicon) copies/µL in qPCR reactions. (B) SARS-CoV-2 N gene damage immediately after UV treatment (Day 0) expressed as log_10_ reduction of N1-2 (long amplicon) copies/µL in qPCR reactions. (C) SARS-CoV-2 N protein concentration measured by ELISA expressed as log_10_ reduction of N protein concentration (pg/mL) in samples immediately after UV treatment (Day 0) and after incubation of samples with host cells (Day 3). SARS-CoV-2 log_10_ reductions of the N1 amplicon, N1-2 amplicon, or N protein versus UV_222_ dose were fit with a linear model forced through the origin at 0 mJ/cm^2^ through responses up to and including 20 mJ/cm^2^ indicated by filled circles. Points not included in models are indicated by hollow circles.

For the long amplicon spanning both the N1 and N2 regions of N gene (CDC 2019), viral RNA damage in response to UV_222_ immediately after treatment was also characterized by exponential decay (Figure 3B). The linear rate constant for LR versus UV_222_ dose was 0.054 ±0.003 cm^2^/mJ (slope ±standard error, R^2^ = 0.94). Compared with the LR rate constant for of SARS-CoV-2 infectivity measured by plaque assay, the LR rate constant of N gene damage measured by N1-2 qPCR was approximately 10-fold lower. This similarity indicates that increasing the amplicon length did not increase the ability to detect gene damage that correlates with loss of viral infectivity. Across all tests, the positive signal for SARS-CoV-2 in the N1-2 assay was 4.6 ±0.1 log_10_ copies/μL in cell cultures infected with untreated virus, undetected in uninfected cell culture supernatants, and 0.8 ±1.4 copies/μL in no template RT-qPCR reaction controls (concentration data and standard curves shown in Supplementary Figures S2 and S3). Because the long amplicon assay was used to investigate potential for improved measurement of disinfection dose response without culture, no Day 3 samples were analyzed.

Although no dose response was observed for LR of the N protein versus UV_222_ dose immediately after treatment for doses up to 40 mJ/cm^2^ (0.002 ±0.001 cm^2^/mJ, slope ±standard error, R^2^ = 0.21), a stronger dose response was observed in Day 3 cell culture supernatants for doses up to 20 mJ/cm^2^ (0.243 ±0.028 cm^2^/mJ, slope ±standard error, R^2^ = 0.21) (Figure 3C). Across all tests, the positive signal for SARS-CoV-2 in the N protein assay was 2.69×10^5^ ±9.83×10^4^ pg/mL in untreated virus samples on Day 0, 1.41×10^8^ ±2.99×10^8^ in Day 3 cell culture supernatants infected with untreated virus, and below detection in uninfected cell culture supernatants (concentration data and standard curves shown in Supplementary Figures S2 and S3). Gene copies/µL for both qPCR assays and protein pg/mL for the ELISA assay are shown for each UV_222_ dose in Supplementary Figure S2 and standard curves for all assays are shown in Supplementary Figure S3.

## DISCUSSION

This study provides the first rigorous UV_222_ dose response kinetics for SARS-CoV-2 in aqueous solution, but there are limitations that must be acknowledged. Most importantly, this study was conducted using virions suspended in aqueous solution. This is only a starting point for quantifying dose response kinetics for airborne virus disinfection that is most relevant for this virus, where many factors such as temperature, humidity, air flow dynamics, and UV reactor specifics will impact dose responses. Previous studies comparing disinfection kinetics of infectious agents in air at increasing relative humidity to those in water^36–41^ indicate that these water dose responses may present a conservative estimate of airborne disinfection kinetics because humidity in many indoor environments is conditioned to reduce infectious agent persistence

One additional limitation of this study related to UV_222_ application in indoor environments is that the disinfection impact of any ozone production by vacuum UV wavelengths potentially emitted by the KrCl excilamp was not measured, but can likely be neglected due to high airflows in the biosafety cabinet and BSL3 facility. The negative air quality impacts and building material degradation by ozone potentially generated by these lamps, and the potential health hazards and building material solarization from wavelengths below 240 nm and the nonzero emission at wavelengths above 240 nm (Supplementary Figure S1), should also be considered when weighing the benefits of reducing infectious disease transmission by UV_222_ for COVID-19 and other infectious diseases.

Considering these limitations, these data provide a strong foundation for future development and application of UV_222_ for reducing airborne viral transmission. UV_222_ is both 4.2 times safer for human exposure (the threshold limit values for human UV exposure are 25 mJ/cm^2^ and 6 mJ/cm^2^ at 222 and 254 nm, respectively^41^) and at least 1.3 times as effective at disinfecting SARS-CoV-2 (the D_90_ we observed for UV_222_ (1.6 mJ/cm^2^) is lower than recently predicted by genomic modeling for UV_254_ (2.15 mJ/cm^2^)^42^). A recent study applying continuous UV_222_ at doses below these threshold limit values to treat other airborne coronaviruses demonstrated multiple logs of inactivation within minutes^43^. This low wavelength advantage for SARS-CoV-2 disinfection is consistent with a study where UV_222_ was more than twice as effective as UV_254_ against MS2 bacteriophage^34^ and with other viral action spectra indicating greater sensitivity at 222 nm than 254 nm^26^. A recent review^44^ predicted the median D_90_ for coronavirus disinfection by UV_254_ to be 3.7 mJ/cm^2^. Our results and these predictions are in general agreement with recent UV_222_ and UV_254_ disinfection studies of SARS-CoV-2 as recently reviewed^41^. However, some of these studies are still in the process of peer review and/or did not use standardized UV disinfection procedures that allow comparisons between experiments and precise quantification of doses. In the only UV_222_ SARS-CoV-2 surface decontamination study to date ^45^, researchers report 0.94 LR after 10 second exposure to 0.1 mW/cm^2^. Although UV dose cannot be calculated for this study in the absence of sample absorbance and differences in experimental setup, these results demonstrate a high degree of susceptibility of SARS-CoV-2 to UV_222_ and generally align with ours.

Considering our data in context of literature, UV_222_ is a promising disinfection method for SARS-CoV-2 in aqueous solution. These infectivity and molecular dose response data could immediately inform measures to prevent transmission by water or wastewater where infectious SARS-CoV-2 and other viruses have been shown to be potentially persistent for days^46,47^. Although tailing was observed in dose responses for molecular assays and may have been contributed from clumping of virus in the protein-laden growth media, viruses were disinfected below detection in plaque assays, indicating that aggregation did not interfere with complete viral inactivation. We did not observe a strong relationship between the kinetics of N gene damage (measured by qPCR with a short and long amplicon) and disinfection, which could reflect that protein damage contributes more to disinfection than genome damage for SARS-CoV-2. One study of MS2 bacteriophage found RNA genome damage to be closely related to and thus contributed to disinfection kinetics^27^, whereas a study of Adenovirus found DNA genome damage not to be closely related to disinfection^48^. This disparity between these viruses with different structures and hosts was further demonstrated when it was shown that protein damage, especially to external capsid proteins, contributes more strongly to UV disinfection of Adenovirus^49^. However, we also did not see a strong association between the kinetics of N protein damage and disinfection. Because we only measured the N protein that closely associates with the viral genome, we may have missed damage to external proteins such as the spike protein which are on the surface to absorb incoming UV radiation and are vital in infection of host cells^50^. Additionally, the confirmation and sequence of the genome and proteins can affect UV genetic damage^51–55^, so the N protein and gene may not be the targets that primarily contribute to disinfection-inducing molecular damage. These factors could explain the weak relationships we observed between disinfection kinetics and N gene damage or N protein damage, and warrant further investigation to unravel the mechanisms of disinfection at this and other UV wavelengths. While these mechanistic complexities remain to be resolved, the disinfection kinetics we report indicate the high degree of susceptibility of SARS-CoV-2 in aqueous solution to UV_222_.

## Supporting information

Cover Letter

## Data Availability

Data for UV dose measurements and calculations, absorbance measurements are available in spreadsheet form upon request. Spreadsheet data for plaque assay results, RT-qPCR assay results, and ELISA assay results are available upon request.

## ACKNOWLEDGEMENTS

This work was supported by funds from The Ohio State University (OSU) Sustainability Institute; OSU Infectious Disease Institute; OSU Department of Civil, Environmental, and Geodetic Engineering (Chair: Dr. Allison MacKay); OSU Department of Microbial Infection & Immunity (Chair: Dr. Eugene Oltz); and National Institutes of Health (U54 CA260582). Anna Herman of AquiSense Technologies shared a list of references to SARS-CoV-2 UV disinfection studies. The UV_222_ light source (USHIO Care222®) was provided by USHIO, Inc. through material transfer agreement 2020-2654 to Hull at OSU.

## CONTENTS

This supplementary information contains Figure S1 describing lamp emission and sample absorbance, Table S1 describing key parameters for UV exposures and dose calculations, Figure S2 standard curves for qPCR and ELISA assays, and Figure S3 molecular assay concentration data for N gene or N protein at each UV dose.

---

**Figure S1:**
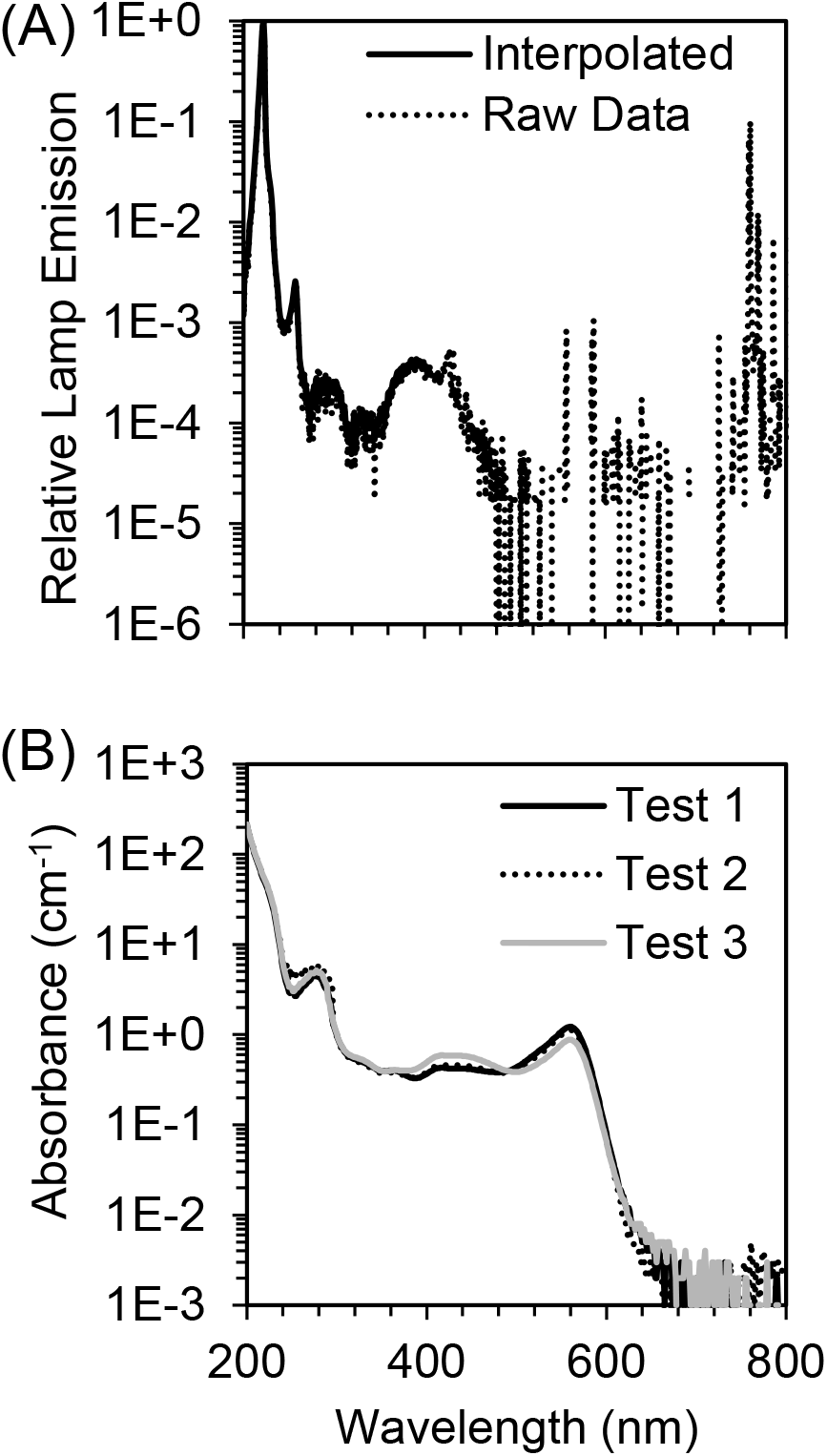
(A) The raw spectral emission from 200 - 300 nm of the filtered excilamp (USHIO Care222^®^) was interpolated and relativized to the peak emission at 222 nm for use in UV dose calculations and plotted on log scale to show orders of magnitude less but non-zero emission at filtered wavelengths > 240 nm. (B) The absorbance spectrum from 200 - 300 nm of SARS-CoV-2 at ∼10^5^ PFU/mL in cDMEM was measured for each of three Tests for use in UV dose calculations.

**Table S1:**
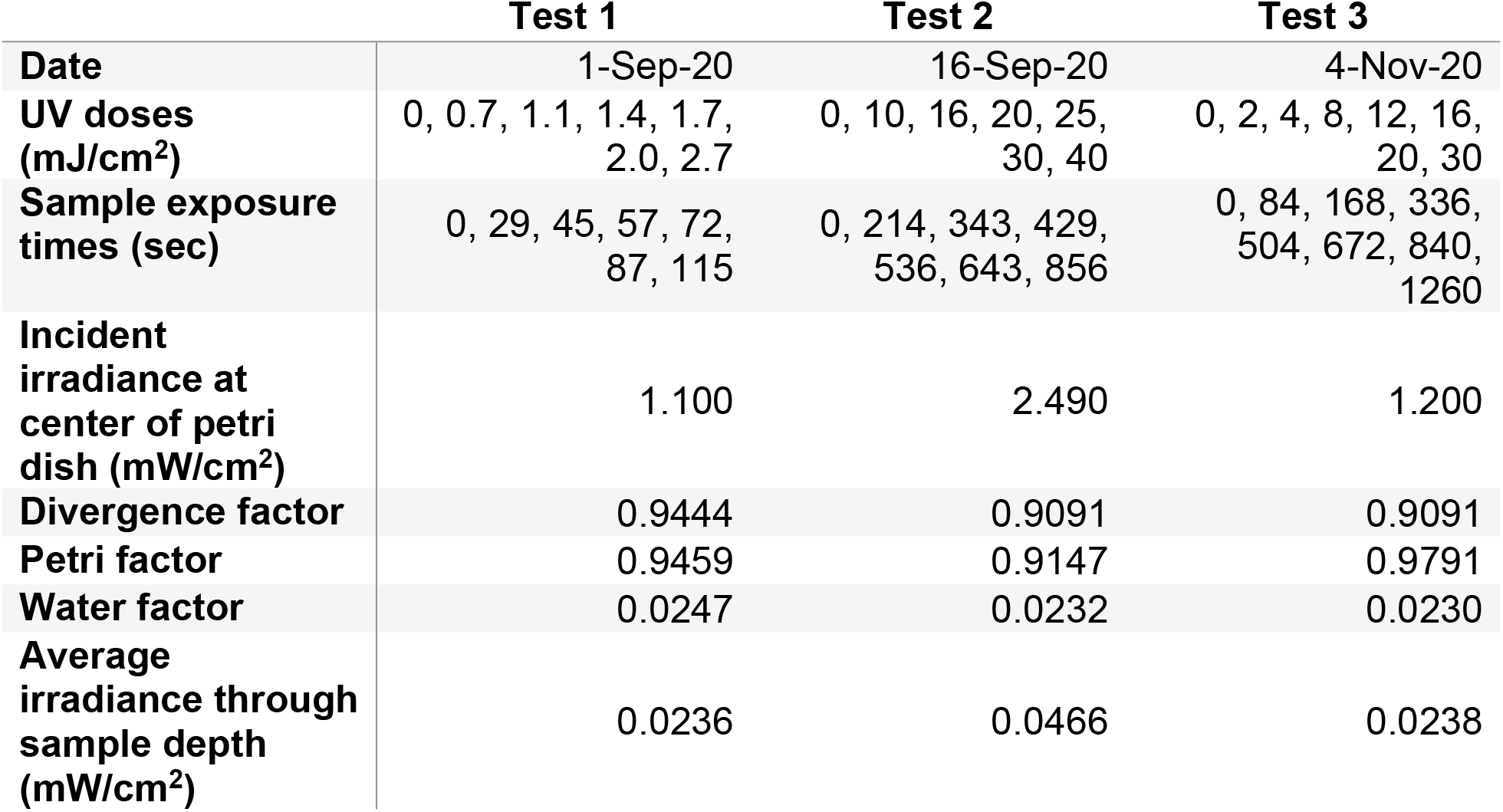
Summary of key UV dose calculation parameters for each independent Test.

|  | **Test 1** | **Test 2** | **Test 3** |
| --- | --- | --- | --- |
| **Date** | 1-Sep-20 | 16-Sep-20 | 4-Nov-20 |
| **UV doses (mJ/cm<sup>2</sup>)** | 0, 0.7, 1.1, 1.4, 1.7, 2.0, 2.7 | 0, 10, 16, 20, 25, 30, 40 | 0, 2, 4, 8, 12, 16, 20, 30 |
| **Sample exposure times (sec)** | 0, 29, 45, 57, 72, 87, 115 | 0, 214, 343, 429, 536, 643, 856 | 0, 84, 168, 336, 504, 672, 840, 1260 |
| **Incident irradiance at center of petri dish (mW/cm<sup>2</sup>)** | 1.100 | 2.490 | 1.200 |
| **Divergence factor** | 0.9444 | 0.9091 | 0.9091 |
| **Petri factor** | 0.9459 | 0.9147 | 0.9791 |
| **Water factor** | 0.0247 | 0.0232 | 0.0230 |
| **Average irradiance through sample depth (mW/cm<sup>2</sup>)** | 0.0236 | 0.0466 | 0.0238 |

**Figure S2:**
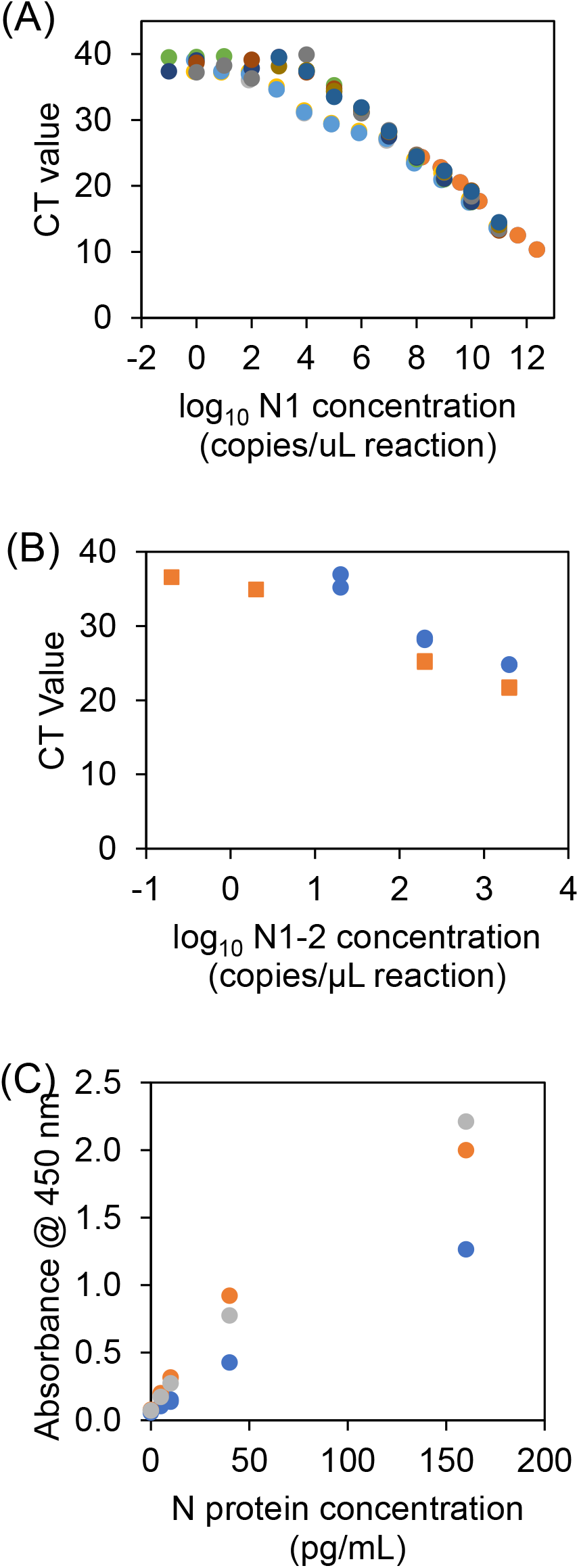
N gene RT-qPCR standard curves gene copies/µL reaction for the (A) short N1 amplicon and (B) long N1-2 amplicon and (C) ELISA N protein standard curve. Colors differentiate individual assay runs.

**Figure S3:**
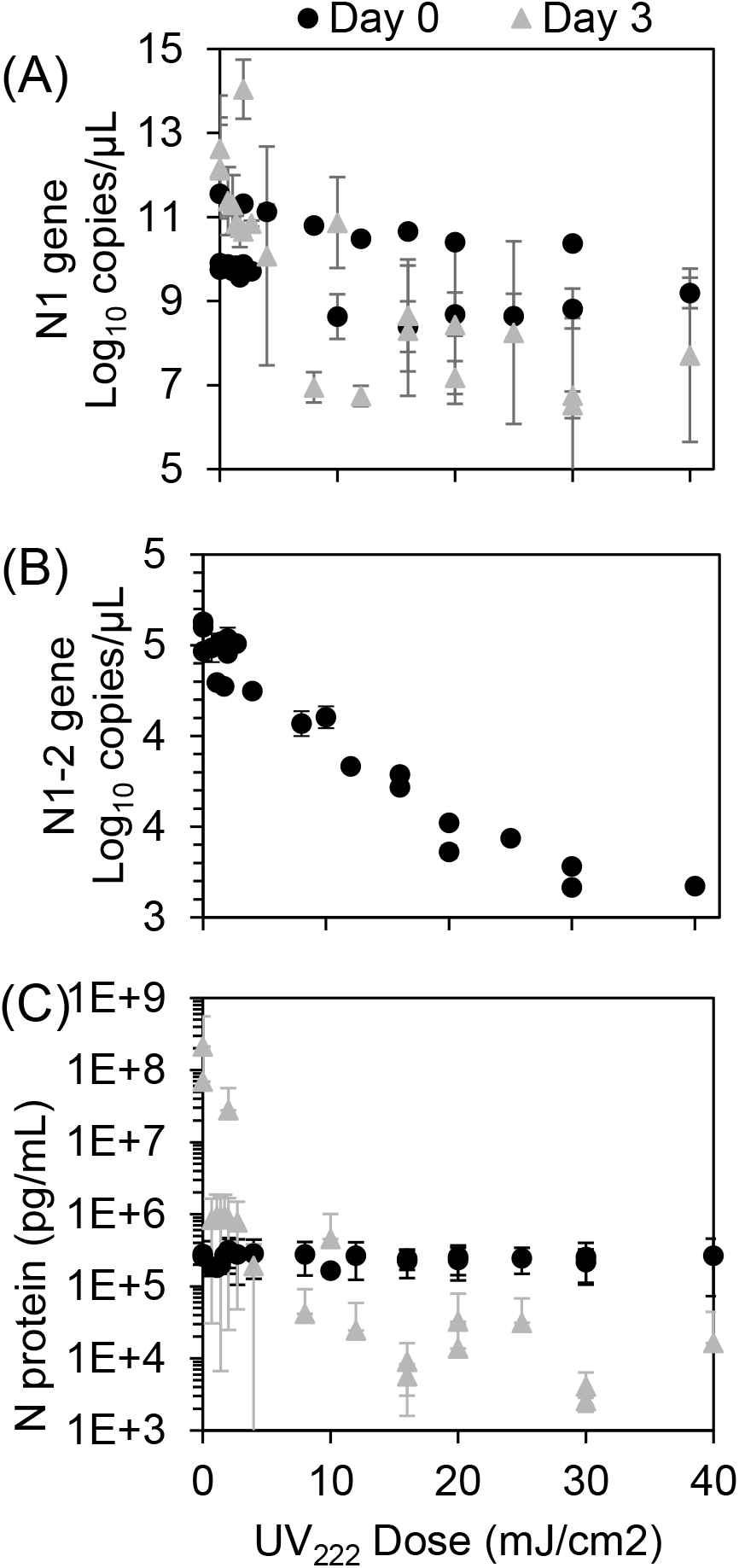
N gene RT-qPCR copies/µL reaction for the (A) short N1 amplicon and (B) long N1-2 amplicon, where error bars represent standard deviation of at least two technical replicates and could include technical replicates averaged across dilutions. (C) N protein ELISA pg/mL, where error bars represent standard deviation of at least two technical replicates and could include technical replicates averaged across dilutions. Day 0 samples were analyzed immediately after UV irradiation, where Day 3 samples were analyzed in culture supernatants after incubation of samples with host cells.

