## Supplementary material for "SARS-CoV-2 disinfection in aqueous solution by UV_222_ from a krypton chlorine excilamp": Cover Letter

THE OHIO STATE UNIVERSITY

College of Engineering

Department of Civil, Environmental and
Geodetic Engineering

ceg.osu.edu

Natalie Hull, Ph.D.
Assistant Professor
Civil, Environmental, and Geodetic Engineering
The Ohio State University
2070 Neil Ave, Hitchcock 417C
Columbus, OH 43210

February 19, 2021

**RE: SARS-CoV-2 disinfection in aqueous solution by $UV_{222}$ from a krypton chlorine excilamp**

Dear Editor,

I am excited to submit for your consideration our article "SARS-CoV-2 disinfection in aqueous solution by $UV_{222}$ from a krypton chlorine excilamp".

Our study is the first rigorous investigation of SARS-CoV-2 disinfection kinetics in aqueous solution. More importantly, this is the first kinetic investigation of SARS-CoV-2 disinfection kinetics in aqueous solution by a UV wavelength (222 nm) emitted by a krypton chlorine excimer lamp. $UV_{222}$ is much less harmful to humans than $UV_{254}$ emitted by conventional mercury-based lamps.

We disinfected SARS-CoV-2 to below our detection limit of 2 PFU/mL at a UV dose of only 8 $mJ/cm^2$, which demonstrated that SARS-CoV-2 is highly susceptible to $UV_{222}$, achieving >4.51 $log_{10}$ (>99.99%) disinfection at 8 $mJ/cm^2$. This low UV dose is only one third of the acceptable threshold dose for human UV exposure at this wavelength. This indicates that very high (>>99.99%) levels of SARS-CoV-2 disinfection could be achieved at UV doses nearing the acceptable threshold dose, or that lower UV doses could be used to minimize risks to human eyes and skin while preventing transmission of airborne virus.

Our results in aqueous solution present a conservative estimate for disinfection kinetics in air where viruses and other microorganisms are more easily inactivated. Taken together with the high $UV_{222}$ susceptibility and improved safety of UV at this far-UV wavelength, this study is important because it supports evidence-based use of $UV_{222}$ as a safe and effective engineering control in public spaces to limit SARS-CoV-2 transmission.

In addition to our culture-based plaque assays, we used RT-qPCR and ELISA to investigate molecular damage induced by $UV_{222}$. We found a statistically significant dose response of RNA damage detected by RT-qPCR using the same assay developed by the CDC and widely used for clinical testing (N1 primer set). This is important because RT-qPCR could be used to measure performance of UV disinfection systems while relying on existing research and testing infrastructure, without the additional biosafety level 3 requirements of culture assays for SARS-CoV-2.

We believe this study is an important step in helping push ahead understanding of how, and how much, SARS-CoV-2 is inactivated in response to $UV_{222}$. This information is critical to informing widespread implementation and monitoring of $UV_{222}$ systems to prevent transmission of SARS-CoV-2 and other emerging and re-emerging infectious diseases.

Sincerely,

Natalie Hull

Natalie Hull
